# *In vivo* amygdala nuclei volumes in schizophrenia and bipolar disorders

**DOI:** 10.1101/2020.09.30.20204602

**Authors:** Claudia Barth, Stener Nerland, Ann-Marie G. de Lange, Laura A. Wortinger, Eva Hilland, Ole A. Andreassen, Kjetil N. Jørgensen, Ingrid Agartz

## Abstract

**Background:** Abnormalities in amygdala volume are well-established in schizophrenia and commonly reported in bipolar disorders. However, the specificity of volumetric differences in individual amygdala nuclei is largely unknown.

**Methods:** Patients with schizophrenia disorders (SCZ, n=452, including schizophrenia, schizoaffective and other psychotic disorders, mean age 30.7±9.2 (SD), females 44.4%), bipolar disorders (BP, n=316, including bipolar I and II, 33.7±11.4, 58.5%) and healthy controls (n=753, 34.1±9.1, 40.9%) underwent T1-weighted magnetic resonance imaging. Total amygdala and nuclei volumes as well as intracranial volume (ICV) were estimated with Freesurfer (v6.0.0). Analysis of covariance and multiple linear regression models, adjusting for age, age^2^, ICV and sex, were fitted to examine diagnostic group and subgroup differences in volume, respectively.

**Results:** Bilateral total amygdala and all nuclei volumes, except the medial and central nuclei, were significantly smaller in patients relative to controls. The largest effect sizes were found for the basal nucleus, accessory basal nucleus and cortico-amygdaloid transition area (partial η2 > 0.02). The diagnostic subgroup analysis showed that reductions in amygdala nuclei volume were most widespread in schizophrenia, with the lateral, cortical, paralaminar and central nuclei being solely reduced in this disorder. The right accessory basal nucleus was marginally smaller in SCZ relative to BP (t = 2.32, p = 0.05).

**Conclusions:** Our study is the first to demonstrate distinct patterns of amygdala nuclei volume reductions in a well-powered sample of patients with schizophrenia and bipolar disorders. Volume differences in the basolateral complex (lateral, basal, accessory basal nuclei) may be putative neuroimaging markers for differentiating schizophrenia and bipolar disorders.

## Introduction

Schizophrenia and bipolar disorders are severe mental health disorders with shared pathophysiological traits along a psychosis continuum (1). Although the neural substrates of the two disorders are still largely unknown, structural brain abnormalities in amygdala volume have been reported in both (2, 3). The amygdala is an almond-shaped brain structure located in the mesiotemporal region of the temporal lobe, adjacent to the hippocampus (4). It is involved in a broad range of complex behaviors, including emotion and threat processing, and the regulation of adaptive behavioral responses (4), known to be affected in both schizophrenia (5) and bipolar disorders (6). Using functional magnetic resonance imaging (MRI), studies on schizophrenia have shown reduced amygdala activation in response to aversive emotional stimuli (7) as well as during facial emotion recognition and evaluation tasks (8, 9). Similarly, abnormal amygdala activity has been observed during emotional processing tasks in patients with bipolar disorders as reviewed by Townsend and Altshuler (10).

On a structural level, there is considerable uncertainty about the specificity and magnitude of volumetric differences in the amygdala in schizophrenia and bipolar disorders. In schizophrenia, whole amygdala volumes showed statistically significant reductions in three large-scale meta-analyses, with low to moderate effect sizes (d≈ 0.2) (2, 3, 11). In bipolar disorders, volumetric reductions have also been shown (12), but they appear to be less pronounced with considerable heterogeneity between studies (13). The causes of the discrepancy in findings are likely multifactorial and complex, and may stem from phenotypic heterogeneity such as differences in disease severity and duration, medication history, and co-morbidities. Particularly medication, such as antipsychotics, antidepressants, antiepileptics, and lithium may impact amygdala volume. For instance, Hartberg and colleagues found lower left amygdala volumes in non-lithium-treated patients, but not in lithium-treated patients (14), and this effect may be dependent on the duration of lithium exposure (15). Differences in neuroimaging data acquisition, amygdala segmentation methods, and analysis approaches, e.g. shape versus volume measures (16, 17), can further affect study outcomes.

The amygdala is not a uniform structure; it rather consists of heterogeneous nuclei with distinct functional correlates (18). Utilizing high-resolution 7T neuroimaging of postmortem brain samples, Saygin and colleagues were able to probabilistically label nine nuclei boundaries, namely the lateral, basal, central, medial, cortical, paralaminar and accessory-basal nuclei, the cortico-amygdaloid transition area, and the anterior amygdaloid area (19). Based on findings from rodent and non-human primate studies, each nucleus seems to serve specific functions via its interconnections with an array of cortical and subcortical structures. The lateral nucleus is the main entry point for sensory information to the amygdala (20): it receives sensory afferent input from the cortex and thalamus, and integrates this information before relaying it to other nuclei. Together with the basal nucleus, the lateral nucleus is an integral part of the threat processing circuitry (21), and postmortem studies indicate that both nuclei may be affected in schizophrenia (22-24).

Recent *in vivo* studies replicated smaller lateral and basal nuclei volumes in first episode psychosis relative to controls (16) and in schizophrenia relative to healthy individuals (25) as well as bipolar patients with psychotic features (17). Volume reductions in the lateral nucleus have also been detected in clinical high-risk individuals (16). Another study in patients with bipolar disorders found significantly smaller volumes in the right basal nucleus, accessory basal nucleus, anterior amygdaloid area, and cortico-amygdaloid transition area relative to controls, but no reductions in the lateral nucleus (26). These findings indicate that reduced lateral nucleus volume may be a risk factor for the development of psychosis, particularly in schizophrenia. Other nuclei may also be reduced in schizophrenia (17, 25), however findings are rather sparse and do not replicate across studies, most likely due to small sample sizes and inclusion of heterogenous diagnostic groups. To our knowledge, no study has systematically investigated amygdala nuclei differences in schizophrenia and bipolar disorders, including schizoaffective disorder, schizophrenia, and other psychotic disorders (OPD), as well as bipolar I and II disorders with or without psychotic features, in a well-powered sample. This is a research gap, as a better understanding of how *in vivo* amygdala nuclei volumes differ in patients with schizophrenia and bipolar disorders may enable more precise and mechanistic theories of amygdala dysfunction in the development of psychosis.

The amygdala is often studied together with the hippocampus (15, 25, 27), as both structures are tightly and reciprocally connected and topologically intertwined (28). Abnormalities in the amygdala-hippocampus complex have been implicated both in schizophrenia (2) and bipolar disorders (13). However, differences in total amygdala and hippocampal volume are rarely directly compared, and it is unclear whether amygdala volume is distinctly or similarly altered relative to hippocampal volume in patients with schizophrenia and bipolar disorders relative to controls.

Here, we examined diagnostic group differences in amygdala nuclei volumes in 768 patients with schizophrenia and bipolar disorders, and 753 healthy controls. Based on previous studies, we expected the strongest volume reductions in patients with schizophrenia, most prominently in the lateral and basal nuclei. We further investigated whether differences in total amygdala volume were comparable to differences in hippocampal volume between diagnostic groups. In exploratory analyses, we also studied potential sources of heterogeneity in volumetric indices based on sex (29), antipsychotic and mood stabilizing medication (14), and psychotic as well as affective symptoms.

## Methods

### Participants

A sample of 753 controls and 768 patients was drawn from the ongoing Thematically-Organized-Psychosis (TOP) study cohort (October 2002 up until January 2018). Patients were recruited from in- and outpatient psychiatric units covering catchment areas in the Oslo region. Healthy controls were randomly drawn from the Norwegian national population register in the same catchment areas. All participants gave written informed consent to participate. The study was approved by the Regional Committee for Research Ethics and the Norwegian Data Inspectorate, and carried out in accordance with the Declaration of Helsinki.

For study inclusion, participants, aged between 18-65 years, were required to have (i) an intelligence quotient (IQ) above 70, (ii) no history of moderate or severe head trauma resulting in hospitalization, and (iii) no neurological disorders or somatic illness which could affect brain structure and function. To obtain a representative clinical sample, patients diagnosed with substance abuse were not excluded, whilst control participants with substance misuse or cannabis use within the last 3 months were.

### Clinical assessment

Clinical diagnoses were established according to the Structured Clinical Interview for Diagnostic and Statistical Manual of Mental Disorders (DSM)-IV axis 1 disorder (SCID-I), module A-E (30). Healthy controls were evaluated with the Primary Care Evaluation of Mental Disorders (Prime-MD) to rule out current or previous psychiatric disorders (31). Presence and severity of psychotic symptoms of patients were assessed using the Positive and Negative Syndrome Scale (PANSS)(32). Affective state were evaluated using the Young Mania Rating Scale (YMRS)(33) and the Inventory for Depressive Symptomatology (IDS)(34). Global Assessment of Function (GAF) scale, split version, was administered to measure general functioning level (35). Clinical characterization was conducted by trained psychologists or psychiatrists.

Based on DSM-IV criteria, the patient sample was diagnosed as follows: (1) *broad schizophrenia spectrum* (N = 452): schizophrenia [DSM-IV 295.1, 295.3, 295.6, 295.9; N = 245], schizophreniform [DSM-IV 295.4; N = 31], schizoaffective [DSM-IV 295.7; N = 59], other psychotic disorders [OPD, DSM-IV 297.1, 298.8, 298.9; N = 117], (2) *bipolar spectrum* (N = 316): bipolar I [DSM-IV 296.0-7; N = 188], bipolar II [DSM-IV 296.89; N = 113], bipolar not otherwise specified [DSM-IV 296.8; N = 15]. The majority of the bipolar patients were investigated in the euthymic phase (87.3 %, i.e. YMRS < 8 and IDS ≤ 13 (36)).

### Medication

Current use of medication among patients, including antipsychotics, antidepressants and antiepileptics, was recorded and converted into Defined Daily Dose (DDD), in accordance with the guidelines from the World Health Organization Collaborating Center for Drug Statistics Methodology. As lithium has a narrow therapeutic window, treatment is routinely optimized by repeated serum concentration measurements to reach optimal exposure for maintenance treatment (37), leaving DDD for lithium less meaningful as a measure of exposure. For this reason, we evaluated lithium user status and serum concentration in blood as a proxy of lithium exposure. Serum blood samples were collected on average 12 or 24 h after last dose according to standard laboratory requirements, and lithium was analyzed by means of commercially available kits using a Cobas Integra 400 plus system (Roche Diagnostics, Rotkreutz, Switzerland, reference range: 0.50 -1-00 mmol/l).

### MRI data acquisition and processing

Participants underwent T1-weighted structural imaging either at 1.5T (2004-2009, N = 745) or at 3T (2011-present, N = 777) scanner. The 1.5T sample was acquired on a Siemens Magnetom Sonata scanner, equipped with a standard head-coil. Two sagittal T1-weighted magnetization prepared rapid acquisition gradient-echo (MPRAGE) volumes were acquired with the Siemens tfl3d1_ns pulse sequence, and averaged to improve signal-to-noise ratio. The acquisition parameters were as follows: echo time (TE) = 3.93 msec, repetition time (TR) = 2730 msec, inversion time (TI) = 1000 msec, flip angle = 7°, field of view = 24 cm, voxel size = 1.33 x 0.94 x 1 mm^3^, number of partitions = 160.

At 3T, T1-weighted 3D Fast Spoiled Gradient Echo (FSPGR) volumes and inversion recovery-prepared 3D gradient recalled echo (BRAVO) volumes were acquired on a General Electric Signa HDxt scanner and a Discovery 750 scanner, respectively. Scanning parameters were as follow: (1) FSPGR (from 2011-2015): TR/TE = 7.8 ms/min, TI = 450 msec, flip angle = 12°, FOV = 256×256 mm^2^, slice thickness = 1.2 mm, acquisition matrix = 256×192, reconstruction matrix = 256×256; (2) BRAVO (from 2015-present): TR/TE/TI 8.16 ms/3.18 ms/450ms, slice thickness = 1.2 mm, reconstruction matrix = 256×256, flip angle = 12°.

T1-weighted MRI images were first processed in FreeSurfer (v6.0.0) using the standard cross-sectional processing stream. Secondly, the module for amygdala nuclei and hippocampal subfields was used to obtain volumes of the left and right amygdala and their respective nuclei, as well as total hippocampus volume and intracranial volume (ICV; estimate based on the Talairach transform). The automated segmentation of the amygdala is based on a probabilistic atlas, created with ultra-high-resolution *ex vivo* MRI data (∼0.1–0.15 mm isotropic), and includes nine subdivisions: the lateral, basal and accessory basal, central, medial, cortical and paralaminar nucleus, the amygdaloid area, and the cortico-amygdaloid transition zone (see Figure 1, for details see (19)). Surface reconstructions were visually inspected, and if necessary, manual editing was performed by trained assistants following standard FreeSurfer procedures (38). To account for the effect of scanner and image acquisition protocols, we used ComBat, a tool adopted from the genomics literature, to remove unwanted variation associated with scanner whilst preserving biological associations in the data (39, 40). ComBat harmonization was performed on amygdala volumes and ICV, with empirical Bayes to leverage information across volumes, and with age, sex and diagnostic group as biological variables of interest.

**Figure 1.**
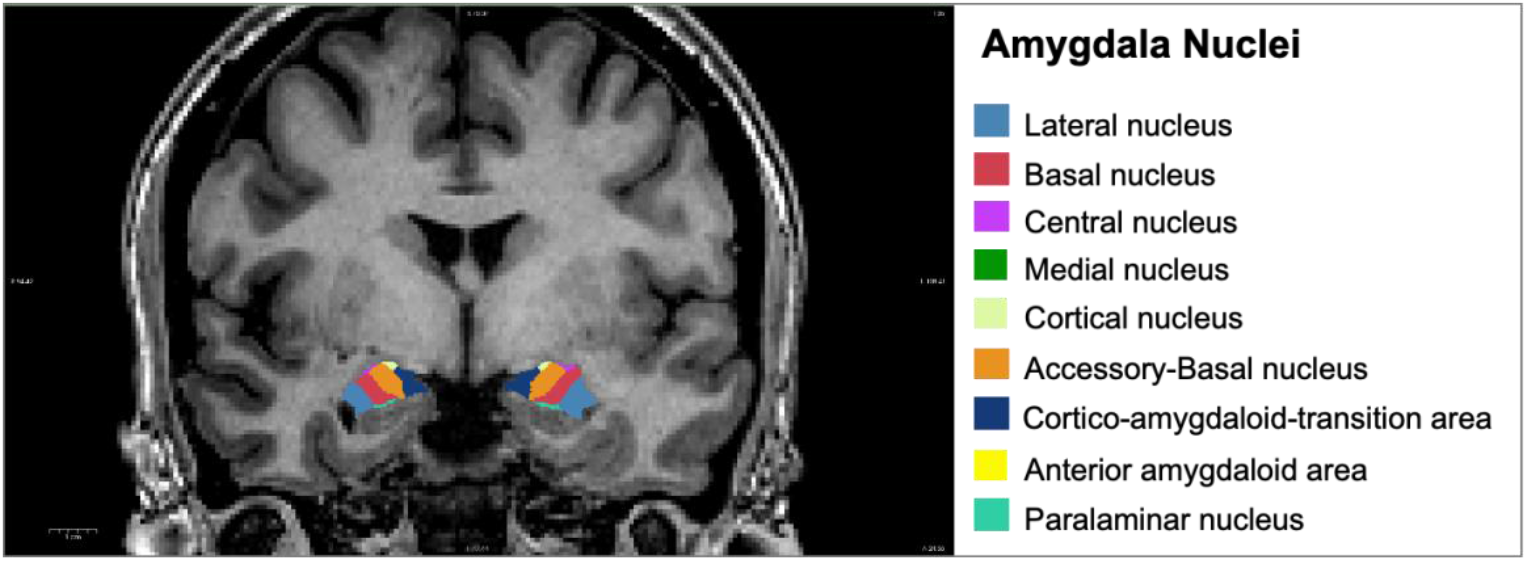
Coronal view of the amygdala nuclei segmentation. Image derived from Freesurfer (v6.0.0).

### Statistical analyses

All statistical tests were conducted in R (v3.5.2; www.r-project.org).

To assess diagnostic group differences in bilateral total amygdala and nuclei volumes across diagnostic groups, we performed analysis of covariance (ANCOVA, type II; *car* package) with volume as dependent variable, diagnostic group and sex as fixed factors and age, age^2^ and ICV as covariates. Age^2^ was added to more accurately model the effect of age, which may have a non-linear relationship with volume (41). *Post hoc* Tukey tests were performed to contrast volume differences between control vs. bipolar disorders, control vs. schizophrenia disorders, bipolar disorders vs. schizophrenia disorders, and females vs. males. Effect sizes were calculated as partial eta-squared (*heplots* package) based on F-statistic. To further explore amygdala volume differences across sex and diagnostic group, we fitted multiple linear models with left/right total amygdala volume as dependent variable and group-by-sex interaction, age, age^2^ and ICV as independent variables.

Multiple linear regression models were also fitted to assess diagnostic subgroup differences (i.e. bipolar I, bipolar II, schizoaffective, schizophrenia, OPD) in total amygdala and nuclei volume, adjusting for age, age^2^, ICV and sex. As we found significantly reduced total amygdala and nuclei volumes in patients relative to controls bilaterally (see Table 2), we combined left and right volumes in the diagnostic subgroup analyses to avoid unnecessary multiple comparisons.

We also evaluated total amygdala volume differences in bipolar I and II patients as well as in bipolar patients with and without psychotic features in additional linear models. The presence of psychotic features was defined based on DSM-IV diagnoses (i.e. 296.44, 296.54, 296.04, 296.64) and a history of psychotic episodes. Diagnostic plots revealed one influential outlier (male bipolar I patient, Cook’s distance > 0.5), which was removed from further analysis. Effect sizes for multiple linear regression results were calculated as Cohen’s d based on t-statistic (*effectsize* package).

As the amygdala is topographically close and strongly, reciprocally connected with the hippocampus, we further examined whether amygdala volume was similarly or differently affected compared to hippocampal volume between diagnostic groups. For this purpose, we fitted separate linear regression models for total amygdala and total hippocampal volume, with the following pairwise group comparisons as independent variables: i) control vs. schizophrenia disorders ii) control vs. bipolar disorders, and iii) schizophrenia disorders vs. bipolar disorders. The models were adjusted for age, age^2^, ICV and sex. Next, we compared the results from the amygdala and hippocampus models for each group comparison using z tests for correlated samples (40), in order to account for the correlations between amygdala and hippocampal volume within each group. The following formula was used, where “m1” and “m2” represent model 1 (amygdala volume) and 2 (hippocampal volume), the β terms represent the beta values from the regression fits, the σ terms represent their errors, and ρ represents the averaged correlation between amygdala and hippocampal volume across the two groups compared:

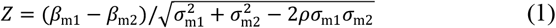

To test for potentially confounding effect of medication on amygdala volumes (dependent variable), additional multiple linear regression models were fitted for each diagnostic group separately. In both schizophrenia and bipolar disorders, the effects of antipsychotics, antidepressants and antiepileptics, measured as DDD (independent variable), were assessed. In bipolar disorders, we further explored the effects of lithium, measured as lithium use status and serum concentration levels, on amygdala volume. The models were adjusted for age, age^2^, ICV, and sex.

To examine the association between amygdala volumes and psychotic symptoms (PANSS subscales in schizophrenia), affective symptoms (YMRS and IDS in bipolar disorder) and general functioning (GAF symptoms/function), additional multiple linear regression models were fitted, accounting for the same covariates as for the medication models.

False discovery rate (FDR) correction was applied to account for multiple comparisons across all volumes tested.

## Results

### Demographic and clinical variables

Sample demographics and clinical characteristics are reported in Table 1. Characteristics of the diagnostic subgroups are summarized in Table S1, supplementary material.

**Table 1.**
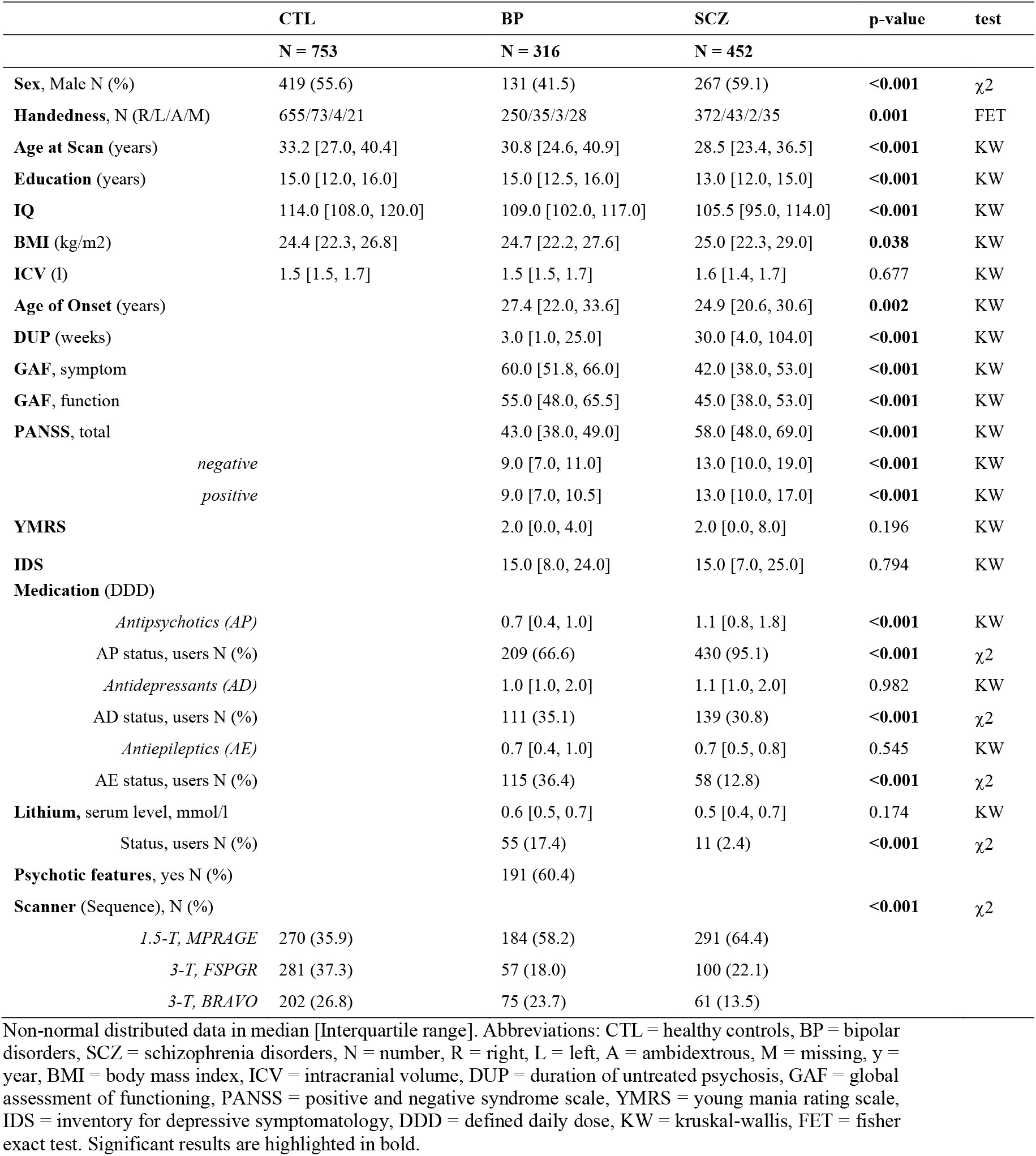
Sample demographics and clinical measures in patients with schizophrenia and bipolar disorders relative to controls.

### Amygdala nuclei volume differences between diagnostic groups

Total amygdala and all nuclei volumes, except the bilateral medial and central nuclei, were significantly lower in patients with schizophrenia and bipolar disorders relative to controls (see Figure 2, Table 2). The strongest effects sizes were found for the basal nucleus, accessory basal nucleus, cortico-amygdaloid transition zone, and the whole amygdala. Post hoc Tukey tests revealed significant differences between healthy controls and patients with bipolar disorders for the bilateral whole amygdala, basal nucleus, accessory basal nucleus, anterior amygdaloid area, cortico-amygdaloid transition area as well as left paralaminar nucleus (see supplementary materials Table S2). Right accessory basal nucleus volume was lower in patients with schizophrenia disorders relative to patients with bipolar disorders (beta = 4.63, standard error (S.E.) = 1.99, t = 2.32, p = 0.053).

**Table 2.**
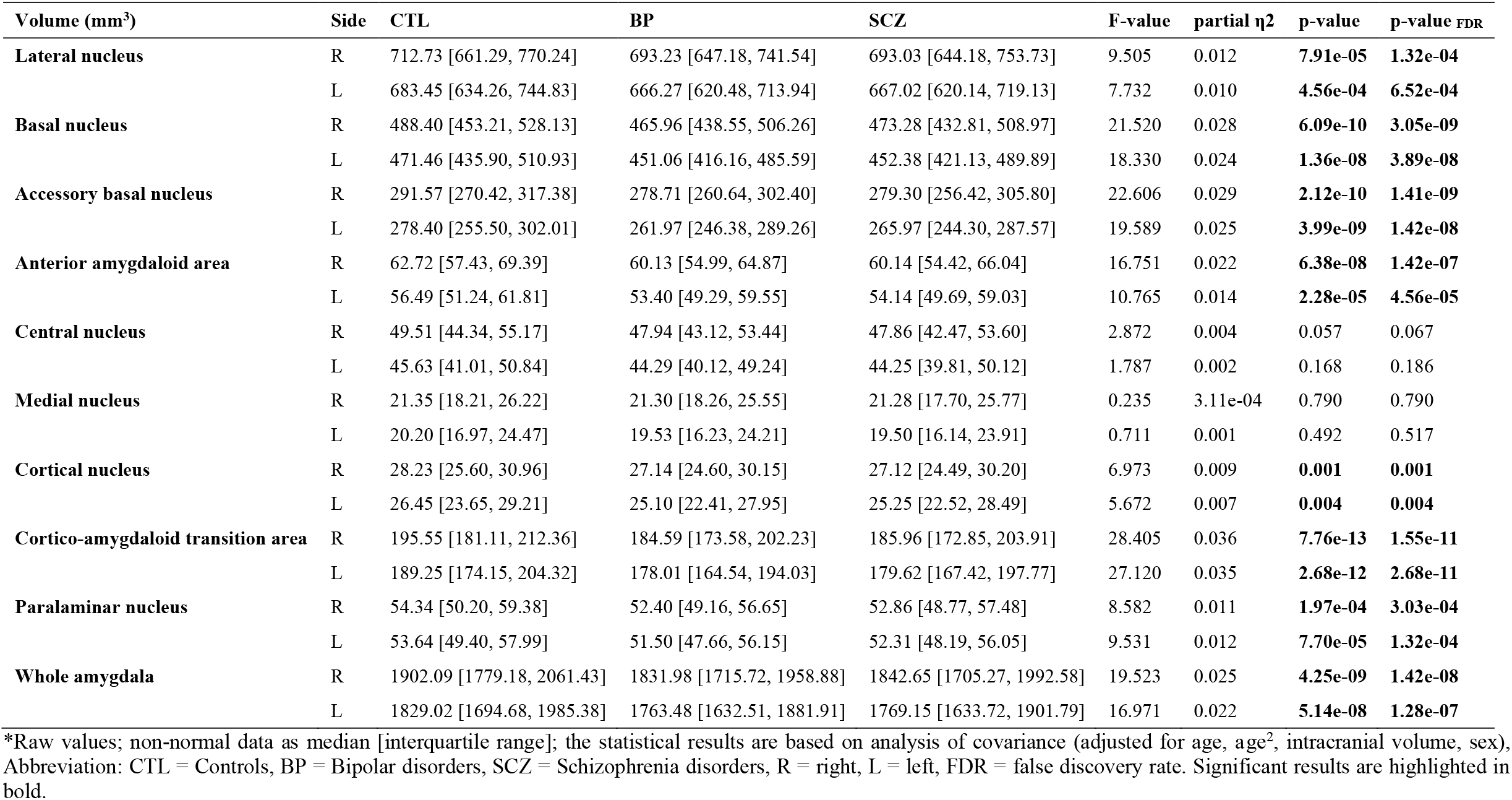
Results of the analysis of covariance of amygdala nuclei in patients with schizophrenia and bipolar disorders relative to healthy controls.

**Figure 2.**
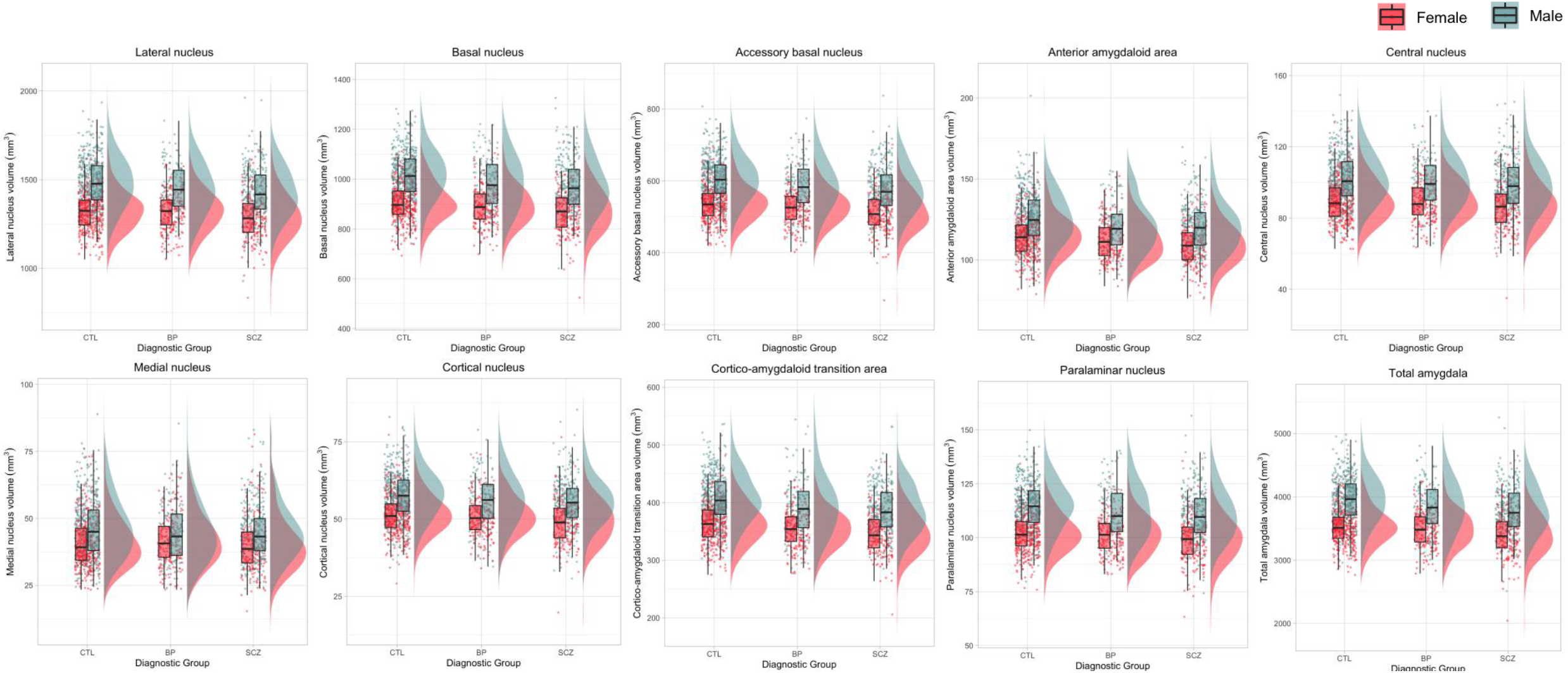
Amygdala nuclei volumes stratified by diagnostic group and sex. Combined for the left and right hemisphere, the volumetric data is displayed as raincloud plots, which combines boxplots, raw data points and the data distributions of data using split-half violins. Females are displayed in red and males in blue. Abbreviation: CTL = healthy controls, BP = bipolar disorders, SCZ = schizophrenia disorders.

Based on the ANCOVA models, we further found a significant effect of sex for all amygdala volumes. Across diagnostic groups, males had higher volumes relative to females (see Figure 2, see supplementary materials Table S2). We found no significant group-by-sex interaction for left (BP vs CTL: beta_Male_ = -26.24, S.E. = 19.32, t = -1.36, p = 0.175; SCZ vs. CTL: beta_Male_ = -18.90, S.E. = 17.10, t = 1.11, p = 0.269) and right total amygdala volume (BP vs CTL: beta_Male_ = -33.28, S.E. = 20.11, t = -1.66, p = 0.098; SCZ vs. CTL: beta_Male_ = -23.05, S.E. = 17.81, t = -1.29, p = 0.196).

The diagnostic subgroup analysis revealed nuclei-specific volume reduction, with schizophrenia showing the most widespread effects (see Figure 3, supplementary materials Table S3). All nuclei, except the bilateral medial nucleus, were significantly smaller in patients with schizophrenia relative to healthy controls. While the basal nucleus, accessory basal nucleus, anterior amygdaloid area and cortico-amygdaloid transition area were smaller in bipolar I, lower nuclei volumes in bipolar II were restricted to the basal nucleus and the cortico-amygdaloid transition area. In OPD, we identified nuclei-specific volume reductions largely overlapping with bipolar I: accessory basal nucleus, anterior amygdaloid area and cortico-amygdaloid transition zone. Patients with bipolar II disorder showed marginally lower total amygdala volume. In patients with schizoaffective disorder, total amygdala volume was not significantly lower relative to controls. Furthermore, we did not find a significant difference in total amygdala volume between neither bipolar I and II patients (beta = 1.86, S.E. = 31.65, t = 0.06, p =0.953), nor between bipolar patients with (n = 191) and without psychotic features (n = 124, beta = 12.16, S.E. = 30.61, t = 0.40, p = 0.691).

**Figure 3.**
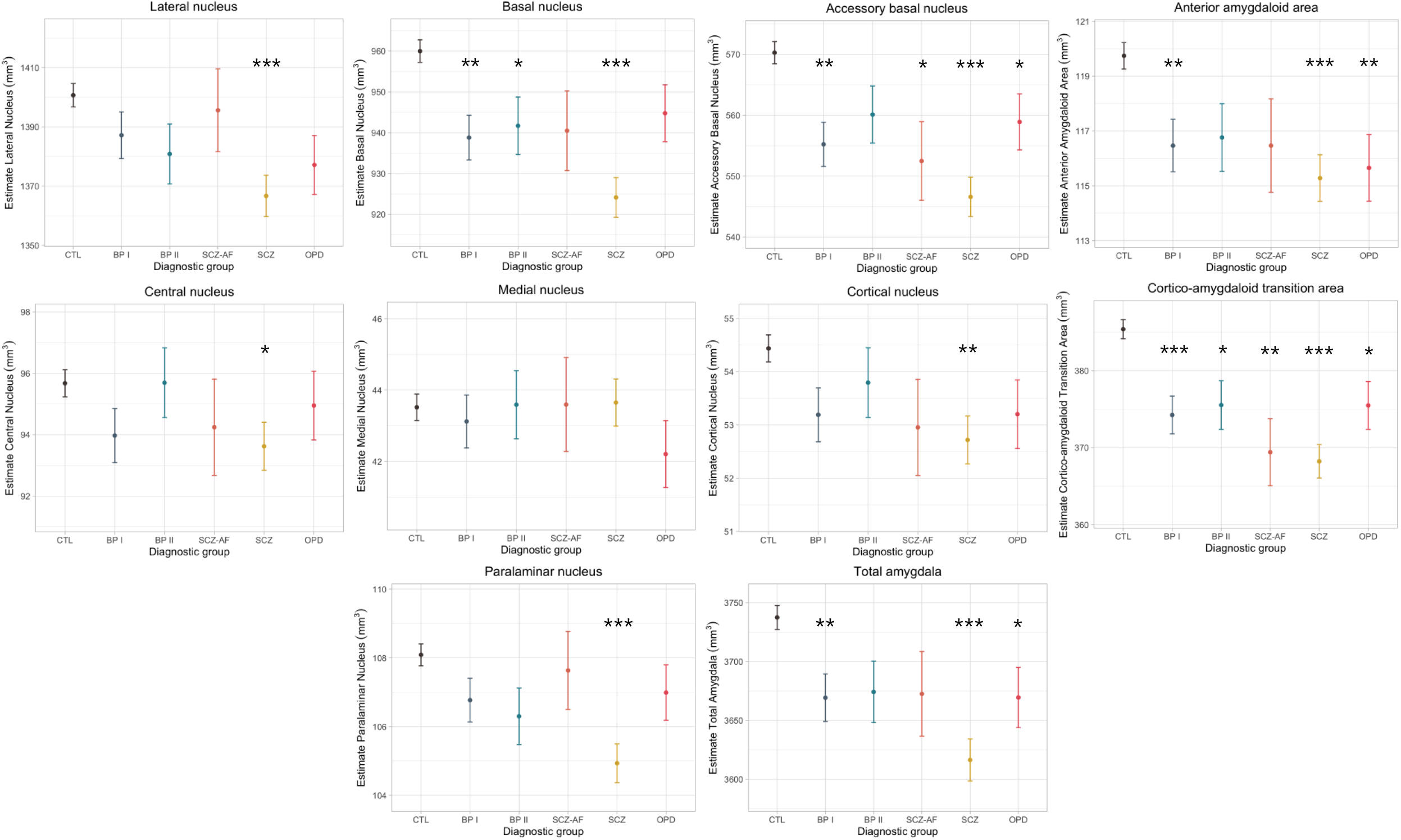
Estimates of total amygdala and nuclei volume stratified by diagnostic subgroup. The model is adjusted for age, age^2^, intracranial volume, and sex. Estimates are displayed with upper and lower confidence intervals. Stars represent significant group difference relative to healthy controls (CTL), after family wise error correction. Significance codes: p < 0.0001 ‘***’; p < 0.001 ‘**’; p > 0.01 ‘*’. BP = bipolar, SCZ = schizophrenia, SCZ-AF = schizoaffective disorder, OPD = other psychotic disorders.

Relative to hippocampal volume, amygdala volume was significantly less reduced in patients compared to healthy controls, while the difference between the schizophrenia disorders and bipolar disorders was not significant (see Table 3). Mean hippocampal volume (mm^3^) for each of the groups was as follows: control 7358.44 ± 695.10 (standard deviation), bipolar disorders 7081.31 ± 661.10, and schizophrenia disorders 7034.74 ± 693.42.

**Table 3.** Pair-wise group differences between amygdala and hippocampus volume based on Z tests (Formula 1).

| Comparison | Amygdala<br>$\beta \pm \text{S.E.}$ | Hippocampus<br>$\beta \pm \text{S.E.}$ | Z-score | P-value | FDR-corrected *p* |
| --- | --- | --- | --- | --- | --- |
| SCZ vs. CTL | -104.83 ± 16.95 | -266.66 ± 30.65 | -7.11 | **1.21e-12** | **3.62e-12** |
| BP vs. CTL | -60.75 ± 19.04 | -153.48 ± 33.78 | -3.60 | **1.22e-04** | **4.80e-04** |
| SCZ vs. BP | -35.31 ± 20.19 | -86.47 ± 38.07 | -1.76 | 0.078 | 0.078 |

Abbreviation: S.E. = standard error, CTL = healthy controls, BP = bipolar disorders, SCZ = schizophrenia disorders, FDR = false discovery rate. Significant results are highlighted in bold.

There were no significant associations between antipsychotic, antidepressant and antiepileptic medication use (DDD) and left or right total amygdala and amygdala nuclei volumes in patients with schizophrenia and bipolar disorders. In patients with bipolar disorders, we found no significant effect of lithium use status or lithium serum concentration on left or right total amygdala and nuclei volume, surviving correction for multiple comparisons. Before FDR correction, however, we found significant positive associations between bilateral cortico-amygdaloid transition area (left: t = 2.25, p = 0.025, p_FDR_ = 0.271; right: t = 2.06, p = 0.041, p_FDR_ = 0.271) as well as bilateral accessory basal nucleus volume (left: t = 2.20, p = 0.029, p_FDR_= 0.271; right: t = 1.93, p = 0.054, p_FDR_ = 0.271) and lithium use status. Similar results were found with lithium serum concentration instead of lithium use status as independent variable.

Left and right total amygdala and nuclei volumes were not associated with positive and negative symptoms in schizophrenia disorders, and not associated with affective symptoms in patients with bipolar disorders. Further, no significant associations were found between general functioning and bilateral amygdala volumes in any of the patient groups.

## Discussion

In the current study, total amygdala and all amygdala nuclei volumes, except the medial and central nuclei, were smaller in patients with schizophrenia and bipolar disorders relative to controls, with bilateral effects. The largest effect sizes were found for the basal nucleus, accessory basal nucleus and cortico-amygdaloid transition area (partial η2 > 0.02). The diagnostic subgroup analysis showed that total amygdala volume was significantly smaller in schizophrenia, bipolar I and OPD, but not in schizoaffective disorder and bipolar II. Reductions in amygdala nuclei volume were most pronounced in schizophrenia: all nuclei, except the medial nucleus, were significantly smaller in patients relative to healthy controls, indicating a more widespread change in amygdala morphology than previously thought.

In schizophrenia, previous postmortem studies (22-24) and more recent *in vivo* findings found smaller nuclei volumes, predominantly, in the basal and lateral nuclei (16) or the right basolateral complex (25), consisting of the lateral, basal and accessory basal nuclei. The lateral nucleus, which extensively projects to the basal and accessory basal nuclei, plays a key role in fear-related responses (21), and lower volume in this subregion has been suggested as a putative biomarker for psychosis risk (16). Here, the lateral nucleus was significantly smaller in schizophrenia, but not in any other diagnostic subgroup, indicating that the lateral nucleus may be particularly implicated in schizophrenia.

However, next to the lateral nucleus, the cortical, paralaminar and central nuclei were also smaller only in schizophrenia. The cortical nucleus is a major target of olfactory projections (42), and the presence of olfactory deficits is well-established in schizophrenia (43). Hence, one might speculate that volume reductions in the cortical nucleus may contribute to this dysfunction. However, we did not find significant volume differences in the medial nucleus between diagnostic groups, a nucleus, which also receives major inputs from the olfactory bulb (44). The paralaminar nucleus projects to the ventral striatum as well as the central nucleus, which is believed to be an important output region for the expression of innate emotional and associated physiological responses (44), and receives inputs from the hippocampus (45). The latter afferents are considered to be involved in contextual fear learning (46). Together with volume reductions in major input regions, such as the lateral nucleus, our findings suggest that both the evaluation of emotional stimuli (input) and the subsequent response (output) are particularly impaired in patients with schizophrenia. In addition, contrary to previous reports, malformations in a network of nuclei rather than volume reductions in selected nuclei such as the lateral nucleus seem to contribute to emotional processing deficits in schizophrenia.

While the lateral nucleus, as part of the basolateral complex, was only reduced in schizophrenia, lower basal and accessory basal nucleus volume was found across diagnostic subgroups. In detail, basal nucleus volume was significantly lower in bipolar I, II, and in schizophrenia, and accessory basal nuclei volume was reduced in all diagnostic subgroups, except bipolar II. Both nuclei play a crucial role in integrating, coordinating, and processing of external sensory information (42, 47). For instance, studies suggest that the basal nucleus decodes emotionally relevant information together with higher-order brain areas (48), to subsequently guide goal-directed behaviors via connections to striatal brain regions (44, 49). Based on these findings, it seems plausible to expect that malformations in basolateral complex may contribute to maladaptive emotional processing and subsequent deficits of adaptive behavior across the schizophrenia-bipolar spectrum. Directly comparing patients with bipolar disorders to patients with schizophrenia disorders, we also found marginally significant volume differences in the right accessory basal nucleus volume, with lower volumes in schizophrenia disorders. One might speculate that exacerbated volume reductions in right accessory basal nucleus, next to other nuclei, may give rise to the more severely disturbed emotional responses, e.g. emotion and face processing (50, 51), in schizophrenia disorders relative to bipolar disorders.

Across all diagnostic subgroups, the cortico-amygdaloid transition area, which is expected to play a crucial role in social communication (47, 52), was consistently smaller relative to controls. Preliminary evidence suggests that the cortico-amygdaloid transition area, as part of the superficial group, participates in the assessment of negative emotions (53), and volume reductions in this sub-region may contribute to the deficits in facial emotion interpretation and social skills observed across the schizophrenia-bipolar spectrum.

The anterior amygdaloid area was also significantly smaller across multiple diagnostic subgroups, namely in schizophrenia, bipolar I and OPD relative to controls. In patients with bipolar disorders, one previous study reported reduced right anterior amygdaloid area volume (26). However, little is known about the connections and functions of the anterior amygdaloid area, making inferences about its implication in these disorders challenging.

Across the bipolar disorder spectrum, fewer amygdala nuclei showed volume reductions in bipolar I than bipolar II when compared to healthy controls. While total amygdala volume as well as basal nucleus, accessory basal nucleus, anterior amygdaloid area and cortico-amygdaloid transition area volume were significantly lower in bipolar I patients relative to controls, volume reductions in bipolar II were limited to the basal nucleus and cortico-amygdaloid transition area. Although amygdala volume differences in bipolar disorders relative to healthy controls have been debated (54), our finding is in line with reports from the ENIGMA bipolar disorder working group showing lower amygdala volume in bipolar I, but not bipolar II (13). More pronounced nuclei volume reductions in bipolar I, similar to schizophrenia, may relate to its more severe symptomatology with a high prevalence of manic episodes and psychotic features. However, we did not find significant total amygdala volume differences between neither bipolar I and II patients nor between bipolar patients with and without psychotic symptoms. The lack of detectable amygdala volume differences between bipolar I (82.4% psychotic) and II (24.0% psychotic) reflects findings from genetic studies, which were also unable to show significant genetic patterns that differentiate these subtypes (55). Although different relative to controls, amygdala morphology appears similar between bipolar I and II, and is likely due to shared genetic underpinnings. Further studies are needed to test whether abnormalities in amygdala structure as well as function may contribute to the distinct clinical manifestation of bipolar subtypes.

The amygdala receives polymodal sensory information from several sources, including the prefrontal cortex, perirhinal cortex, and hippocampus (42). Projections between the amygdala nuclei and hippocampus are particularly strong and reciprocal (28). This interdependency raises the question whether the observed morphological alterations in amygdala volume translate to similar or different changes in hippocampal volume across diagnostic groups. Here, we found that total amygdala volume was significantly less reduced than the total hippocampus volume in patients compared to healthy controls, while the difference between the schizophrenia and bipolar disorders was not significant. These results indicate that hippocampal morphology, relative to the amygdala, seems more affected in patients with schizophrenia and bipolar disorders, but to a similar extent. Efferents from the basolateral complex to the hippocampus and other brain regions are believed to be glutamatergic (56), and idiopathic psychoses, including schizophrenia and mood disorders with psychotic features, may arise from abnormal glutamatergic neurotransmission in the hippocampus (57). Abnormalities in amygdala morphology and projections to the hippocampus might exacerbate hippocampal dysfunction, reflected in greater volume reductions detected by MRI. However, due to reciprocal projections, the effect could also be reversed: higher hippocampal volume reductions may lead to morphological abnormalities in the amygdala. Future studies may examine whether lower nuclei volumes are associated with decreased connectivity between the amygdala and the hippocampus as well as other brain regions important for emotional processing in schizophrenia and bipolar disorders.

We found a significant effect of sex for all amygdala nuclei volumes, with higher volumes in males relative to females, after correcting for intracranial volume. This finding is in line with results from a recent cohort study, including 2,838 healthy adults (29). However, we did not find a sex-by-diagnostic group interaction, indicating that the effect of sex may be uniform across diagnostic groups.

Although medication, including antidepressants and lithium, have been shown to increase amygdala volume (14, 58), we found no association between exposure to psychotropic drugs or mood stabilizers and volume in schizophrenia and bipolar disorders. However, putative medication effects are likely multifactorial and driven by overall medication history and duration of exposure. Longitudinal studies are needed to shed light on the neuroplastic effects of specific medications on amygdala volume in psychosis disorders. Furthermore, total amygdala volume was not significantly associated with any measure of symptom severity, including positive and negative symptoms in schizophrenia disorders and affective symptoms in bipolar disorders. However, these results are in line with previous reports not showing any associations between amygdala volume and symptom measures (59-61). The lack of significant associations between morphology and proxies of burden of illness is unclear, but may relate to diagnostic issues such as marked heterogeneity among psychotic disorders, imprecise measurement tools, variability of symptom states over time, or currently unknown confounders.

The major strength of the current study is the large sample size with a largely balanced sex distribution, and careful and detailed assessment of clinical characteristics of patients with schizophrenia and bipolar disorders, including medication use and symptom profiles. However, the cross-sectional nature of the presented data does not enable causal inference, and longitudinal studies are needed to determine the timing of morphologically changes in amygdala nuclei in patients. Furthermore, as the amygdala is a small subcortical structure, parsing this region into nuclei using MRI is challenging. Different manual and automatic segmentation approaches have been developed, however, the method deployed here is the first to use ultra-high-resolution *ex vivo* MRI data (∼ 0.1–0.15 mm isotropic) (19), which implies higher sensitivity of our study to detect nuclei-specific structural abnormalities in the amygdala. Although advantageous, this also poses limitations as the position of the internal boundaries between the nuclei are probabilistically labelled based on the *ex vivo* training data, and the volumes of the nuclei must thus be interpreted with caution. In addition, although we harmonized amygdala volumes and ICV across different scanners using ComBat, and automatic volumetry has been shown to be reliable in multicenter MRI studies (62), residual effects of scanner may still be present.

In summary, our study is the first to highlight distinct patterns of amygdala nuclei volume reductions in a well-powered sample of patients with schizophrenia and bipolar disorders. Further research is needed to replicate our findings, parse out the clinical significance of amygdala nuclei reductions, and assess the propensity of nuclei-specific malformations to serve as putative biomarkers to distinguish psychiatric disorders.

## Data Availability

We made use of publicly available resources for processing the imaging data and for conducting the statistical analyses. The clinical and neuroimaging data can be made available upon request.

## Acknowledgments

We thank the study participants, and the clinicians as well as research assistants involved in recruitment and assessment at the Norwegian Centre for Mental Disorders (NORMENT). This work was supported by the Research Council of Norway (grant numbers 223273, 250358, 286838) and the K. G. Jebsen Foundation (grant number SKGJ-MED-008).

